# Impact of the Clinical Trials Act on noncommercial clinical research in Japan: An interrupted time-series analysis

**DOI:** 10.1101/2021.01.29.21250729

**Authors:** Ikuyo Tsutsumi, Yusuke Tsutsumi, Chikashi Yoshida, Takuya Komeno, Yuichi Imanaka

## Abstract

**Background:** The number of new noncommercial clinical studies conducted in Japan declined within the first year of the implementation of the Clinical Trials Act (CTA) on April 1, 2018. This study aimed to examine the impact of the CTA’s enforcement on the number of new noncommercial clinical studies registered in the Japanese Clinical Trial Registry.

**Methods:** An interrupted time-series design was used in the analysis, which was conducted for the period of April 2015 to March 2019. We collected data for trials registered in the Clinical Trial Registry, managed by the University Hospital Medical Information Network.

**Results:** In total, 35,811 studies were registered in the registry; of these, 16,455 fulfilled the eligibility criteria. The difference in the trend of monthly number of new trials after CTA enforcement decreased significantly by 15.0 trials (95% CI, −18.7 to −11.3), and the level decreased by 40.8 (95% CI, −68.2 to −13.3) from the pre-enforcement to the post-enforcement period. Multigroup analyses indicated that the act exerted a significant effect on the trend of new clinical trials, particularly those with smaller sample sizes, interventional study designs, and nonprofit funding sponsors.

**Conclusions:** The number of Japanese noncommercial clinical studies declined significantly following implementation of the CTA. It is necessary to establish a system to promote clinical studies in Japan while ensuring transparency and safety.

## Introduction

Recently, various regulations and laws have been developed for clinical research worldwide, because research misconduct and inappropriate relationships between pharmaceutical companies and researchers have become serious problems. Such “research scandals” have recently become important ethical issues in Japan as well. It was exposed that data falsification and conflicts of interest occurred in research conducted in various fields, resulting in papers being withdrawn in several clinical studies and serious confusion in clinical practice [1]. Research misconduct can impair data accuracy and cause disadvantages to participants and those who would benefit from the results.

Clinical studies requiring approval for drugs and medical devices are legally regulated by the International Conference on Harmonization Guidelines Defining Good Clinical Practice ordinance in Japan. However, there are no legal regulations governing noncommercial clinical research receiving funds or benefits from manufacturers and using unapproved or off-label use drugs or medical devices. To promote the conduct of clinical research by ensuring trust in clinical research and thereby improve public health and hygiene, the Clinical Trials Act (CTA) was established by the Ministry of Health, Labour and Welfare (MHLW) [2] in Japan and enforced since April 1, 2018. It aimed to define procedures for the conduct of clinical research, appropriate provision of the management of reviews by certified review board (CRB), and systems for disclosure of information regarding funding or other benefits for clinical research [3]. Since the enforcement of the CTA, such clinical trials have become subject to legal control as “specified clinical trials” [3,4]. The CTA requires 1) contracts and disclosure of information regarding research funding; 2) review of the implementation plan and adverse events by a CRB authorized by the MHLW; 3) compliance with implementation standards for monitoring, conflict of interest management, and record preservation; and 4) disclosure of the implementation plan to the MHLW.

While legal regulations for clinical research have been implemented in various countries to ensure transparency, overregulation can sometimes limit clinical research, particularly that of a noncommercial nature [5,6]. In the European Union (EU), the number of studies submitted for research grants or ethical review has declined by 30% to 50%, while that of noncommercial studies has decreased from 40% to 14% since Directive 2001/20/EC was adopted in April 2001 and launched in May 2004 [7,8]. Similarly, in Japan, there is the concern that the new law could reduce the number of studies conducted at research institutes lacking financial support, but this has not been examined previously. This study aimed to clarify the association between the enforcement of the CTA and the number of studies newly registered in the Japanese Clinical Trial Registry, using an interrupted time-series analysis design.

## Materials and methods

### Data source

We collected data from the Clinical Trials Registry managed by the University Hospital Medical Information Network (UMIN-CTR) [9], which is part of the Japan Primary Registries Network (JPRN). The JPRN consists of the following 3 clinical research registration agencies: UMIN-CTR; Japan Pharmaceutical Information Center Clinical Trial Information [10]; and the Clinical Trials Registry operated by the Center for Clinical Trials of the Japan Medical Association [11]. The JPRN is a clinical trial registry that meets the criteria of the International Committee of Medical Journal Editors and was authorized by the World Health Organization (WHO) Primary Registry on August 16, 2008 [12]. Academic clinical studies are registered in the UMIN-CTR. However, pharmaceutical company-led commercial clinical trials are registered in the Japan Pharmaceutical Information Center Clinical Trial Information, and physician-led commercial and medical device clinical trials are registered at the Center for Clinical Trials, Japan Medical Association. Additionally, the newly established clinical research database, the Japan Registry of Clinical Trials (jRCT), was added to the JRPN and approved by the WHO Primary Registry on December 5, 2018. The UMIN-CTR provides open .csv files, including daily snapshots of studies registered in the database. We downloaded the file from the UMIN-CTR website on April 1, 2019 [9].

### Outcomes

The primary outcome was the change in the trend, defined as the difference in changes (slope) in the monthly number of new clinical studies, before and after the enforcement of the CTA. The secondary outcome was the change in the level of the monthly number of new clinical studies, defined as the difference in the monthly number of new clinical studies from the end of the pre-CTA period to the period immediately following the enforcement of the CTA. Additionally, the study focused on differences in the effects of the CTA on the following factors: sample size (≤100/>100), study objectives (malignancy/nonmalignancy), study design (interventional/noninterventional), and type of funding sponsor (for-profit/nonprofit).

### Data selection

The inclusion criterion was an anticipated trial start date between April 1, 2015 and March 31, 2019. The exclusion criteria were the exclusion of Japan from the study region and commercial research requiring Investigational New Drug applications to the MHLW, because these studies were applicable to the International Conference on Harmonization Guidelines Defining Good Clinical Practice, rather than the CTA.

### Statistical analysis

Descriptive analyses were performed based on the baseline characteristics of studies before and after the enforcement of the CTA. Continuous variables are presented as medians (interquartile ranges [IQRs]), and categorical variables as frequencies and percentages. A Wilcoxon rank-sum test was performed to compare continuous variables, and Pearson’s chi-squared test was used in between-group comparisons of categorical or binary variables. An interrupted time-series analysis (ITSA) design [13,14] was used to assess the association between the enforcement of the CTA and changes in the trends and levels of the monthly number of new studies. The intervention of interest was the enforcement of the CTA. The first month of the intervention period was set as April 2018, and the analysis period lasted for 48 months, from April 2015 to March 2019. To create the time-series dataset, the aggregate number of studies for each month from April 2015 to March 2019 was tabulated according to the anticipated trial start date. We performed ITSA using two ordinary least-squares regression-based approaches [15].

The following regression equation was used in the single-group analysis [15–18]:

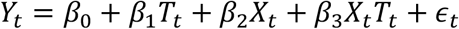

The following regression equation was used in the multigroup analysis [14–17]:

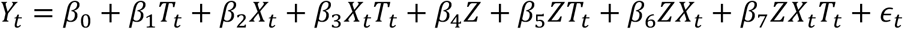

*Y*_*t*_ represents the monthly number of studies measured at time point *t*, and *T*_*t*_ represents the time since April 2015. *X*_*t*_ is a dummy variable representing the enforcement of the CTA (e.g., pre-CTA period was 0, otherwise 1). In the single-group analysis, β_0_ represents the number of studies in the first month of the study (i.e., April 2015). β_1_ represents the slope of the monthly number of studies (trend) before the implementation of the CTA. β_2_ represents the change in the monthly number of studies from the end of the pre-CTA period (level change) to the period immediately following the enforcement of the CTA. β_3_ indicates the slope change following the enforcement of the CTA. In the multigroup analysis, β_0_ to β_3_ represents the value for the control group, and β_4_ to β_7_ represents the value for comparison group. β_4_ represents the difference in the numbers of studies during the first month of the study between the control group and comparison group (difference in level) prior to enforcement of the CTA. β_5_ represents the difference in the slopes of the monthly number of studies between the control group and comparison group (difference in trend) prior to enforcement of the CTA. β_6_ represents the difference in levels immediately following enforcement of the CTA between the control group and comparison group. β_7_ represents the difference in slopes (trends) in the pre- and post-CTA periods between the control group and comparison group. Calendar month was included as a dummy variable, to account for seasonality. Newey-West standard errors were used to deal with autocorrelation and possible heteroskedasticity [15]. All statistical tests were two-sided and the significance level was set at 5%. All analyses were performed using Stata SE version 14.2 (Stata Corp, College Station, TX, USA). The requirement for ethics committee approval was waived because all the data are publicly available online and comprise only aggregate values, without any personally identifiable information.

## Results

Fig 1 shows the flow chart for data selection. We downloaded all data for 35,811 studies in the UMIN-CTR on April 1, 2019. The 502 studies that were not conducted in Japan, and the 57 that involved Investigational New Drug applications to the MHLW, were excluded according to the exclusion criteria. Additionally, 18,797 studies in which the anticipated trial start date was not between April 1, 2015 and March 31, 2019 were excluded from the analysis. Therefore, 16,455 studies ultimately met the selection criteria.

**Fig 1.**
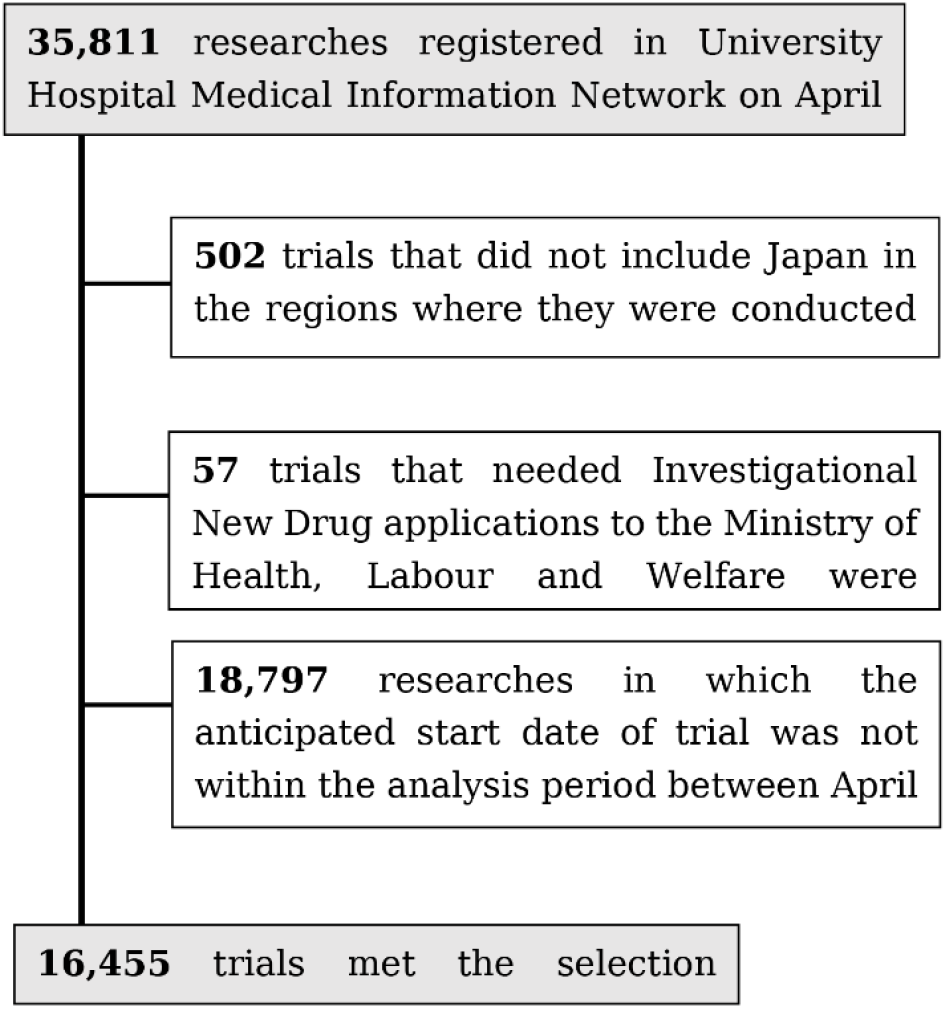
Flow diagram of data selection. UMIN-CTR: University Hospital Medical Information Network; MHLW: Ministry of Health, Labour and Welfare.

Table 1 shows the baseline characteristics of studies initiated between April 2015 and March 2019. The proportion of interventional studies conducted after the enforcement of the CTA was lower relative to that of those conducted before the enforcement of the CTA (from 70.8% to 66.6%, *p* < .001). Regarding disease classification, the proportion of studies involving internal medicine and surgery conducted after the enforcement of the CTA was lower relative to that of those conducted before (from 43.0% to 40.0%, *p* = .002 and from 12.6% to 9.3%, respectively). Moreover, the proportion of studies involving malignancy decreased after the law was enforced (from 24.4% to 21.7%, *p* < .001). Contrarily, the proportion of studies involving healthy people increased after the enforcement (from 22.6% to 29.0%, *p* < .001). As regards types of funding organizations, the most common was self-funded, followed by for-profit organizations and Japanese Governmental offices, both before and after the enforcement of the CTA.

**Table 1.**
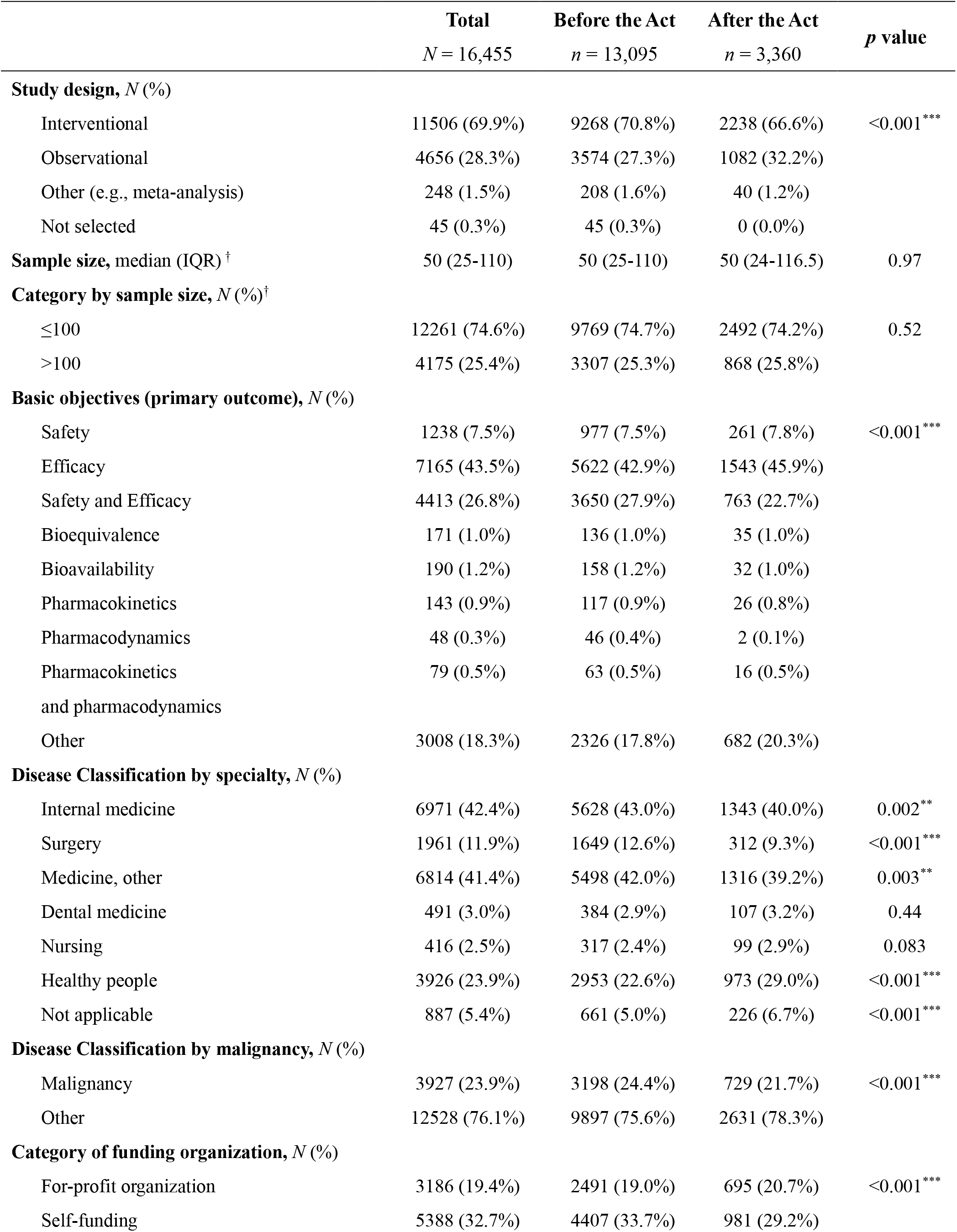
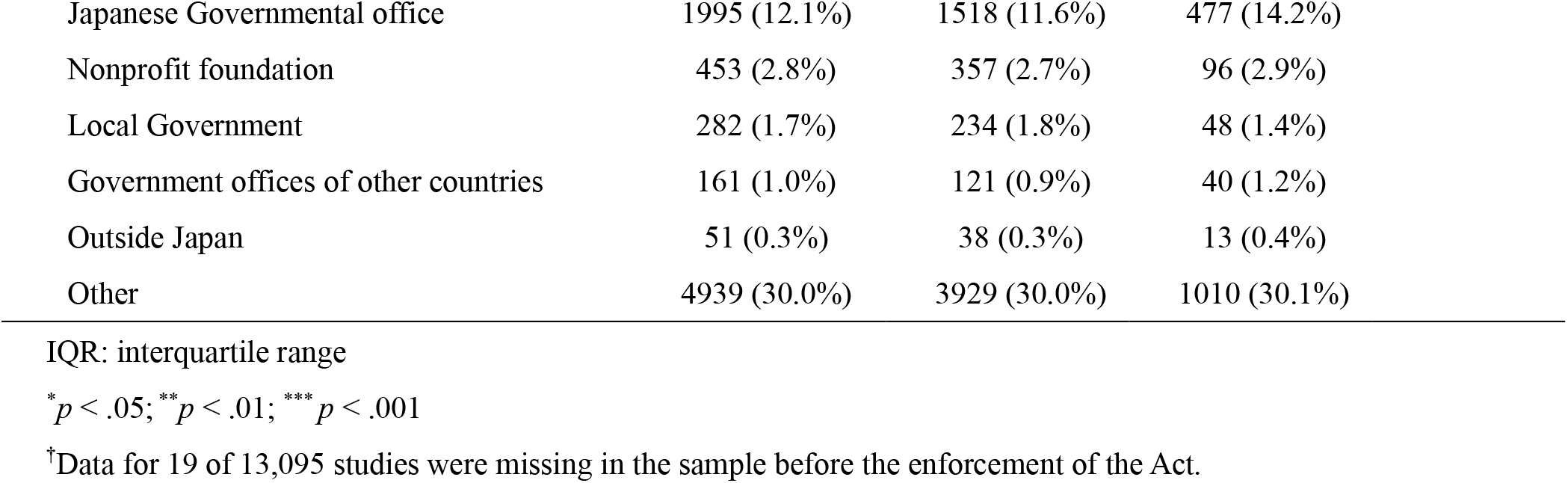
Baseline characteristics of trials that began between April 2015 and March 2019.

|  | Total<br>*N* = 16,455 | Before the Act<br>*n* = 13,095 | After the Act<br>*n* = 3,360 | *p* value |
| --- | --- | --- | --- | --- |
| **Study design, *N* (%)** |  |  |  |  |
| Interventional | 11506 (69.9%) | 9268 (70.8%) | 2238 (66.6%) | <0.001*** |
| Observational | 4656 (28.3%) | 3574 (27.3%) | 1082 (32.2%) |  |
| Other (e.g., meta-analysis) | 248 (1.5%) | 208 (1.6%) | 40 (1.2%) |  |
| Not selected | 45 (0.3%) | 45 (0.3%) | 0 (0.0%) |  |
| **Sample size, median (IQR) <sup>†</sup>** | 50 (25-110) | 50 (25-110) | 50 (24-116.5) | 0.97 |
| **Category by sample size, *N* (%) <sup>†</sup>** |  |  |  |  |
| ≤100 | 12261 (74.6%) | 9769 (74.7%) | 2492 (74.2%) | 0.52 |
| >100 | 4175 (25.4%) | 3307 (25.3%) | 868 (25.8%) |  |
| **Basic objectives (primary outcome), *N* (%)** |  |  |  |  |
| Safety | 1238 (7.5%) | 977 (7.5%) | 261 (7.8%) | <0.001*** |
| Efficacy | 7165 (43.5%) | 5622 (42.9%) | 1543 (45.9%) |  |
| Safety and Efficacy | 4413 (26.8%) | 3650 (27.9%) | 763 (22.7%) |  |
| Bioequivalence | 171 (1.0%) | 136 (1.0%) | 35 (1.0%) |  |
| Bioavailability | 190 (1.2%) | 158 (1.2%) | 32 (1.0%) |  |
| Pharmacokinetics | 143 (0.9%) | 117 (0.9%) | 26 (0.8%) |  |
| Pharmacodynamics | 48 (0.3%) | 46 (0.4%) | 2 (0.1%) |  |
| Pharmacokinetics<br>and pharmacodynamics | 79 (0.5%) | 63 (0.5%) | 16 (0.5%) |  |
| Other | 3008 (18.3%) | 2326 (17.8%) | 682 (20.3%) |  |
| **Disease Classification by specialty, *N* (%)** |  |  |  |  |
| Internal medicine | 6971 (42.4%) | 5628 (43.0%) | 1343 (40.0%) | 0.002** |
| Surgery | 1961 (11.9%) | 1649 (12.6%) | 312 (9.3%) | <0.001*** |
| Medicine, other | 6814 (41.4%) | 5498 (42.0%) | 1316 (39.2%) | 0.003** |
| Dental medicine | 491 (3.0%) | 384 (2.9%) | 107 (3.2%) | 0.44 |
| Nursing | 416 (2.5%) | 317 (2.4%) | 99 (2.9%) | 0.083 |
| Healthy people | 3926 (23.9%) | 2953 (22.6%) | 973 (29.0%) | <0.001*** |
| Not applicable | 887 (5.4%) | 661 (5.0%) | 226 (6.7%) | <0.001*** |
| **Disease Classification by malignancy, *N* (%)** |  |  |  |  |
| Malignancy | 3927 (23.9%) | 3198 (24.4%) | 729 (21.7%) | <0.001*** |
| Other | 12528 (76.1%) | 9897 (75.6%) | 2631 (78.3%) |  |
| **Category of funding organization, *N* (%)** |  |  |  |  |
| For-profit organization | 3186 (19.4%) | 2491 (19.0%) | 695 (20.7%) | <0.001*** |
| Self-funding | 5388 (32.7%) | 4407 (33.7%) | 981 (29.2%) |  |

|  |  |  |  |
| --- | --- | --- | --- |
| Japanese Governmental office | 1995 (12.1%) | 1518 (11.6%) | 477 (14.2%) |
| Nonprofit foundation | 453 (2.8%) | 357 (2.7%) | 96 (2.9%) |
| Local Government | 282 (1.7%) | 234 (1.8%) | 48 (1.4%) |
| Government offices of other countries | 161 (1.0%) | 121 (0.9%) | 40 (1.2%) |
| Outside Japan | 51 (0.3%) | 38 (0.3%) | 13 (0.4%) |
| Other | 4939 (30.0%) | 3929 (30.0%) | 1010 (30.1%) |

IQR: interquartile range
\* $p < .05$ ; \*\* $p < .01$ ; \*\*\* $p < .001$
<sup>†</sup>Data for 19 of 13,095 studies were missing in the sample before the enforcement of the Act.

Fig 2 shows the results of the single-group ITSA for the monthly number of new studies. During the pre-enforcement period, the trend in the number of new studies increased by 1.64 (95% CI: 0.71 to 2.57, *p* = .001) each month. On the other hand, during the post-CTA period, this trend was expected to decline significantly, by 13.3 (95% CI: -17.1 to -9.63, *p* < .001) every month. The difference in trends before and after the enforcement of the CTA was -15.0 (95% CI: -18.7 to -11.3, *p* < .001). Furthermore, there was a significant decline in levels (−40.8, 95%CI: -68.2 to -13.3, *p* = .005) after the enforcement.

**Fig 2.**
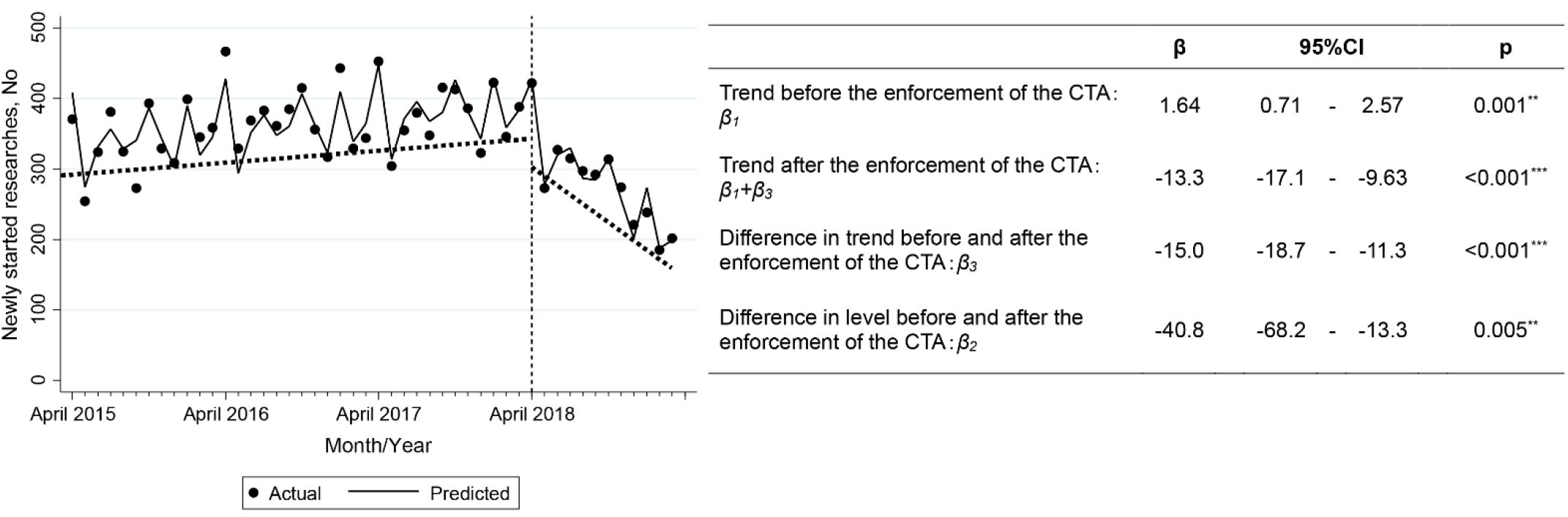
Results of the single-group ITSA for monthly number of new studies. The points on the figure represent the actual monthly number of studies. The solid lines indicate the predicted monthly number of studies adjusted by calendar month. Dotted lines represent trends (slopes) in monthly numbers of studies. Level change after the enforcement of the Clinical Trials Act (CTA) is defined as the difference from the end of the dotted line during the pre-Act period to the starting point of the dotted line during the post-Act period. ITSA: interrupted time-series analysis; CI: confidence interval.

Fig 3 represents the results of the multigroup ITSA examining the effects of the CTA on various factors such as sample size, study design, and type of funding sponsor. As shown in Fig 3a, the pre-CTA trend for studies with sample sizes >100 increased by 0.53 (95% CI: 0.22 to 0.84, *p* = .001) monthly, which did not show a significant difference from that of studies with sample sizes ≤100 (0.65, 95% CI: -0.29 to 1.58, *p* = .17). In contrast, the post-CTA trend for studies with sample sizes >100 decreased by 2.99 (95% CI: -5.33 to -0.65, *p* = .013) monthly, while studies with sample sizes ≤100 showed a significant difference from that of studies with sample sizes >100 (−7.33, 95% CI: -10.6 to -4.11, *p* < .001). There was no significant difference in level changes between studies with sample sizes ≤100 and >100 during the period immediately following the enforcement of the CTA (−15.8, 95% CI: -45.4 to 13.8, *p* = .29).

**Fig 3.**
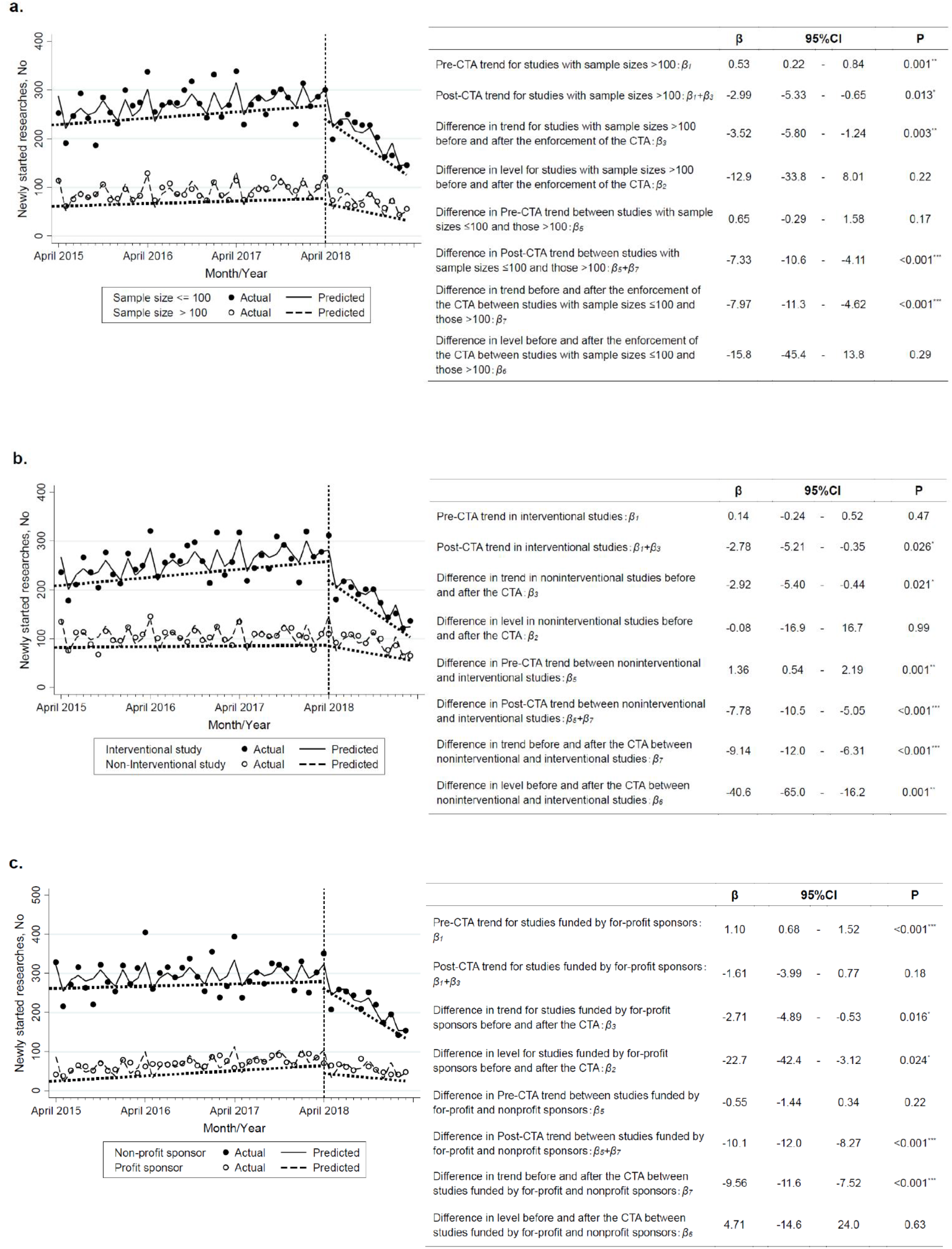
Results of the multigroup ITSA for monthly number of new studies. (a) ITSA according to sample size. (b) ITSA according to study design. (c) ITSA according to type of funding sponsor. The points on the graph represent the actual monthly number of studies. Solid lines indicate the predicted monthly number of studies adjusted by calendar month. Dotted lines represent trends (slopes) in monthly number of studies. Level change after the enforcement of the Clinical Trials Act (CTA) is defined as the difference from the end of the dotted line during pre-Act period to the starting point of the dotted line during post-Act period. ITSA: interrupted time-series analysis; CI: confidence interval.

Moreover, Fig 3b shows the effects of the CTA according to study design. The pre-CTA trend in noninterventional studies did not increase significantly (0.14, 95% CI: -0.24 to 0.52, *p* = .47), while interventional studies increased significantly by 1.36 (95% CI: 0.54 to 2.19, *p* = .001) per month. The post-CTA trend for noninterventional studies decreased by 2.78 (95% CI: -5.21 to -0.35, *p* = .026) per month, while interventional studies showed a significant difference from that for noninterventional studies (−7.78, 95% CI: -10.5 to -5.05, *p* < .001). There was a significant difference in level changes between interventional and noninterventional studies during the period immediately following the enforcement of the CTA (−40.6, 95% CI: -65.0 to -16.2, *p* = .001).

Furthermore, Fig 3c represents the results of the ITSA according to the type of funding sponsor. The pre-CTA trend for studies funded by for-profit sponsors increased at a rate of 1.10 (95% CI: 0.68 to 1.52, *p* < .001) per month, which did not differ significantly from studies funded by nonprofit sponsors (−0.55, 95% CI: -1.44 to 0.34, *p* = .22). In contrast, the post-CTA trend for studies funded by for-profit sponsors decreased at a rate of 1.61 (95% CI: -3.99 to 0.77, *p* = .18) per month, while studies funded by nonprofit sponsors showed a significant difference from that for studies funded by for-profit sponsors (−10.1, 95% CI: -12.0 to -8.27, *p* < .001). There was no significant difference in level changes between studies funded by for- and nonprofit sponsors during the period immediately following the enforcement of the CTA (4.71, 95% CI: -14.6 to 24.0, *p* = .63).

## Discussion

The results of the single-group ITSA showed that the total number of new studies declined significantly in both trends and levels following the enforcement of the CTA in April 2018. The current data indicated that the enforcement of the CTA exerted a strong negative effect on the number of new clinical studies. The analysis of various factors, using the multigroup ITSA method, showed that the trend decreased significantly for all types of research after the new legal regulation, especially those with smaller sample sizes, interventional study designs, and nonprofit funding sponsors. The result suggests that enforcement of the CTA affected studies with limited human resources and financial support in particular. Prior to implementation, there were no legal restrictions in Japan on noncommercial clinical research. After introduction of the new law, CRB authorized by the MHLW reviewed research plans, adverse event reports, and the exact status of conflicts of interest of all physicians involved in the study [2–4]. These improvements seek to increase the transparency of the procedures involved in clinical research and flow of expenses, which contribute to the prevention of research misconduct. However, substantial research funding is needed to disburse expensive commission fees for review by a CRB and management costs, including personnel expenses for following requirements mandated by the CTA, as compared to the period prior its introduction. Therefore, it can be difficult for researchers who do not have sufficient human resources and funds to conduct new clinical research.

Since 2001, each country in the EU has developed its own directives for clinical research to maintain the quality of research based on Directive 2001/20/EC. Thus, in most countries, the number of noncommercial studies has decreased, particularly those funded by nonprofit sponsors [19–21], because regulatory bodies impose highly demanding stipulations and expensive fees for the submission of research to ethics committees. There are similar concerns in Japan that the CTA may also overly regulate clinical researches regardless of the research subject and magnitude of the risk to participants. Only a limited number of organizations with accessible labor force and adequate financial resources could conduct clinical studies mostly with larger sample sizes, which may limit the scopes of study domains. Furthermore, it may cause a decline in the number of new researchers who challenge innovative clinical research. It is essential to develop a system in which physicians can obtain appropriate support to conduct the studies. We may be able to follow the model developed in Italy, where the number of clinical studies increased after the introduction of legal regulation [21]. This is apparently because Italy implemented policy changes that include waiving of ethical review fees, prompt approval by ethics review boards, financial support for research expenses or management, and alleviation of regulations on investigator-driven study for noncommercial research [21–23].

This study was subject to a few limitations. First, it did not include jRCT data analyzed in this study. All noncommercial researches certified as a “specified clinical trial” are required to register with the jRCT since the enforcement of the CTA. However, there were only six new studies registered in the jRCT but not in UMIN-CTR between April 1, 2015 and March 31, 2019 as reported in the WHO International Clinical Trials Registry Platform. This suggests that exclusion of the jRCT data has little impact on the current results. Second, we could analyze the data for only one year after the enforcement of the CTA. It may be essential to carry out medium- and long-term analysis for examining the impact of the CTA on clinical studies in Japan. Nevertheless, the current analysis contributes to identify the key issues of Japanese clinical studies under the new legal regulation to solve them quickly. Finally, it is difficult to assess whether the introduction of CTA could really reduce clinical studies exclusively with inadequate transparency and reliability, which is the original purpose of the legislation, despite the decrease in newly initiated trials found in this study. Further study from an alternative perspective may be needed to clarify the issue.

## Conclusions

The number of noncommercial clinical studies has decreased 1 year after the implementation of the CTA in Japan. This could lead to the decline of the credibility of Japanese clinical research. Establishing a new system to promote clinical research in Japan while ensuring research transparency and safety is vital.

## Data Availability

The datasets generated and/or analyzed during the current study are available in the UMIN-CTR repository, https://www.umin.ac.jp/ctr/csvdata.html.

## Acknowledgments

We would like to thank Editage (www.editage.cn) for English language editing.

